# Accelerated long-term forgetting in drug-resistant focal epilepsy: Insight from a prospective controlled study using a multimodal associative memory paradigm

**DOI:** 10.64898/2026.01.08.26343621

**Authors:** Victoria Guinet, Emmanuelle Dantony, Hélène Catenoix, Mikaïl Nourredine, Samuel Garcia, Sébastien Boulogne, Mathilde Leclercq, Charlotte Le Guen, Nicolas Fourcaud-Trocmé, Pascal Roy, Nadine Ravel, Sylvain Rheims

## Abstract

**Background:** Episodic memory complaints are frequently reported by people with epilepsy (PWE), and often contrast with normal performance on standard neuropsychological assessment using short retention intervals (20-30 minutes). This discrepancy is consistent with the concept of accelerated long-term forgetting (ALF). However, the objective assessment of ALF remains challenging in clinical practice, and the underlying mechanisms, whether related to encoding, early or late consolidation, are still debated. This study aimed to evaluate a specifically designed associative memory task to detect episodic memory impairment and ALF in PWE who show normal performance on standard neuropsychological assessment.

**Methods:** In a prospective non-randomised controlled study, 40 PWE and 40 healthy controls (HC) completed an associative memory task comprising the encoding of 56 abstract word-naturalistic scene pairs, followed by retrieval at 30min and 72h. At each delay, both recollection and recognition were assessed on complementary halves of the encoded material. During encoding, participants had to provide for each pair a subjective judgement about the congruence between the word and the scene. The primary endpoint was recollection-based memory loss 72h after encoding in PWE compared to HC, analysed using a logistic mixed-effects model.

**Results:** Memory significantly declined 72h after encoding, with a lower recollection score at 72h than at 30min in both groups (PWE: OR [95%CI]: 0.31 [0.26;0.38]; HC: OR [95%CI]: 0.21 [0.17;0.26]). At 30-min recollection, PWE were less likely to perform correctly than HC (OR [95%CI]: 0.49 [0.38;0.63]) and despite a smaller memory loss over time, PWE continued to perform significantly worse than HC at 72h (OR [95%CI]: 0.73 [0.58;0.91]. No association was found between memory performance and epilepsy-related clinical features. A three-way interaction was observed between the congruence judgement, group and time (p=0.03), with PWE recovering normal recollection performance at 30 min for pairs judged as highly congruent during encoding.

**Discussion:** The results suggest that ALF in epilepsy may reflect disruption occurring early in the consolidation process, potentially during encoding itself and underline the importance of associative, multimodal episodic memory tasks to better characterise memory impairment in PWE.

## INTRODUCTION

People with drug-resistant focal epilepsy (PWE) frequently report episodic memory impairment^1,2^ that often contrasts with normal performance on standard neuropsychological measures^3,4^. This discrepancy is commonly interpreted as accelerated long-term forgetting (ALF), a cognitive phenomenon characterised by apparently intact learning but disproportionately rapid forgetting over hours to weeks^5,6^. First documented in temporal lobe epilepsy (TLE), particularly in transient epileptic amnesia, ALF has since been described across a range of neurological conditions^7–12^, suggesting shared underlying mechanisms. Although not disease-specific, ALF appears to be particularly prevalent in TLE^13^, given the anatomical overlap between memory networks and epileptogenic sites, as well as potential aggravating factors such as interictal epileptiform activity during sleep^14^.

From a theoretical perspective, ALF is commonly conceptualised as a consolidation disruption, however, whether it arises from early or later stages of this process remains debated^15^. Given the various methodologies used to detect it^16,17^, evidence further suggests that it may reflect disturbances at encoding^18,19^, a stage poorly explored at the behavioural level. This strongly highlights the need to refine current assessment methods, which are often central to patient management and particularly critical in the pre-surgical evaluation of TLE. Although efforts have been made to harmonise neuropsychological assessment procedures in epilepsy surgery^20,21^, findings from research on memory consolidation are still struggling to be translated into clinical practice. Thus, exploring episodic memory under rigorous controlled conditions is challenging. This largely reflects the complexity and dynamic nature of this function which relies on a distributed cerebral network that predominantly involves medial temporal lobes structures^22^. Within this network, the hippocampus plays a key role from the initial stages of memory formation by binding together perceptual components of an experience to their contextual features (e.g., spatial, temporal, emotional). Throughout memory consolidation, the hippocampus maintains close interactions with neocortical regions. It remains strongly involved in the retrieval of both recent and remote personal events, as such recollection requires the reconstruction of perceptual and contextual details associated with the original experience^23–25^. Consequently, episodic memory is inherently associative, and its assessment requires materials that strongly engage this dimension to detect disturbances within the hippocampal-neocortical network supporting episodic memory formation and consolidation.

In this context, we aimed to evaluate whether a specifically designed associative memory task could detect episodic memory impairment and measure ALF in PWE who exhibit normal results on a comprehensive neuropsychological assessment.

## METHODS

### 1. Study design

The EPIMNESIE study (NCT04924933) was a prospective non-randomised controlled study conducted between 2022 and 2025 at the Adult Epilepsy Department of Hospices Civils de Lyon, Lyon, France. Its objective was to evaluate the diagnostic value of an original associative memory task in people with epilepsy (PWE) compared with healthy controls (HC). The task comprised an encoding phase followed by two recall sessions at 30 minutes and 72 hours, each including both recollection and recognition tasks. The outcomes of the memory task were examined with respect to recollection performance on the one hand, and recognition performance on the other. Given that recollection performance represents the most clinically relevant measure, the present analysis focuses on recollection-related outcomes, whereas recognition-related ones will be presented separately in a dedicated analysis. The EPIMNESIE study complied with the TREND statement^26^ (supplemental Text S1).

### 2. Standards Protocol Approvals, Registrations, and Patient Consents

EPMNESIE was approved by ethics committee (CPP OUEST IV, 59/21_2, 09/2021) and all participants, PWE and HC, gave their written informed consent.

### 3. Participants

#### 3.1 Inclusion criteria

All participants were French-native speakers, had no comprehension, reading or sensory impairment, and were not likely to be pregnant.

PWE were screened during a routine medical visit by one of the investigators experts in epilepsy (HeCa, SB or SR). The inclusion visit was scheduled once the neuropsychologist investigator (VG) confirmed that the following inclusion criteria were met: (i) adult (≥ 18 years old) with a diagnosis of drug-resistant focal epilepsy according to the International League against Epilepsy criteria^27^ and (ii) who proved normal results in a recent (< 2years) comprehensive neuropsychological assessment performed for clinical purpose and fulfilling the French consensus for neuropsychological assessment procedure (NAP) in epilepsy surgery^20^. In addition to screening for potential psychiatric comorbidities, this routine evaluation assessed several domains such as language, visuospatial abilities, memory, executive functioning, attention, processing speed and intellectual efficiency using tests listed in the Supplemental Text S2. Tests results were considered normal if superior to a -1.5 SD threshold. Only patients who demonstrated normal performance in all memory tests were eligible for inclusion. Mild impairments in other cognitive dimensions were tolerated, provided that these domains remained normal according to the IC-CoDE criteria^28^, with the same threshold applied. Patients were not included if they showed significant depressive symptoms (French version of the Neurological Disorders Depression Inventory for Epilepsy^29^ – NDDIE score >15), had undergone epilepsy surgery, or had experienced a seizure within the 12 hours preceding the test sessions.

HC were recruited in parallel by circulating a call for volunteers using a specific announcement detailing the study objectives, within hospital and university networks. Inclusion criteria for HC were: (i) no history of neurological or psychiatric disorder; (ii) normal scores on the Montreal Cognitive Assessment^30^ (MoCA, score >27) and on the Matrix Reasoning subtest from the Wechsler Adult Intelligence Scale^31^ (WAIS-IV, scaled score >5). HC were not included if they showed significant depressive or anxiety symptoms on the Hospital Anxiety and Depression Scale (HADS^32^, score ≥8 in either dimension) or reported any episodic memory complaint.

#### 3.2 Demographic and clinical variables

Demographic data were collected at inclusion for both groups (age, sex, years of education, handedness). Additional clinical data comprised, for PWE: NDDIE score at inclusion and epilepsy characteristics, including duration of the disease, lobar localisation and lateralisation of the epileptogenic zone (EZ), MRI data, seizure frequency and number of focal to bilateral tonic-clonic seizures over the last 3 months, current antiseizure medications (ASM) and other treatments. The number of seizures occurring during the study, particularly during the 72-h interval, was also collected; for HC: HADS, MoCA and Matrix Reasoning scores.

### 4. Associative memory task

#### 4.1 Design of the behavioural task

A computerised associative memory task was specifically designed by two investigators, experts in cognitive neuroscience (NR) and neuropsychology of epilepsy (VG). It was implemented in Python (SG) and a script was developed to present the stimuli and automatically record participants’ responses, which were all provided via a standard computer keyboard. The task comprised three test sessions (Figure 1): (i) an encoding of 56 pairs composed of an abstract word and a photo of a naturalistic scene depicting various environments; (ii) a 30-min recall during which both a recollection and a recognition tasks were used; (iii) a 72-h recall using the same procedure as the 30-min recall. To avoid potential practice effects, the material was not repeated from one recall to the next. Thus, the 30-min recall was based on half of the pairs presented during encoding (n=28), whereas the 72-h recall was based on the remaining half (n=28), to which the participants had therefore not been re-exposed during the delay. Examples were provided before all sessions to ensure that the instructions were properly understood and the task feasible. For recollection and recognition tasks, the participants were not informed about the different types of distractors described below. To reduce fatigue and allow for optimal level of attention, breaks were included during the recall sessions. A fixed inter-stimulus interval of 1500ms (black screen followed by a fixation cross) was used in all sessions. The administration of the associative memory task was performed by the same investigator (VG) for all participants.

**Figure 1.**
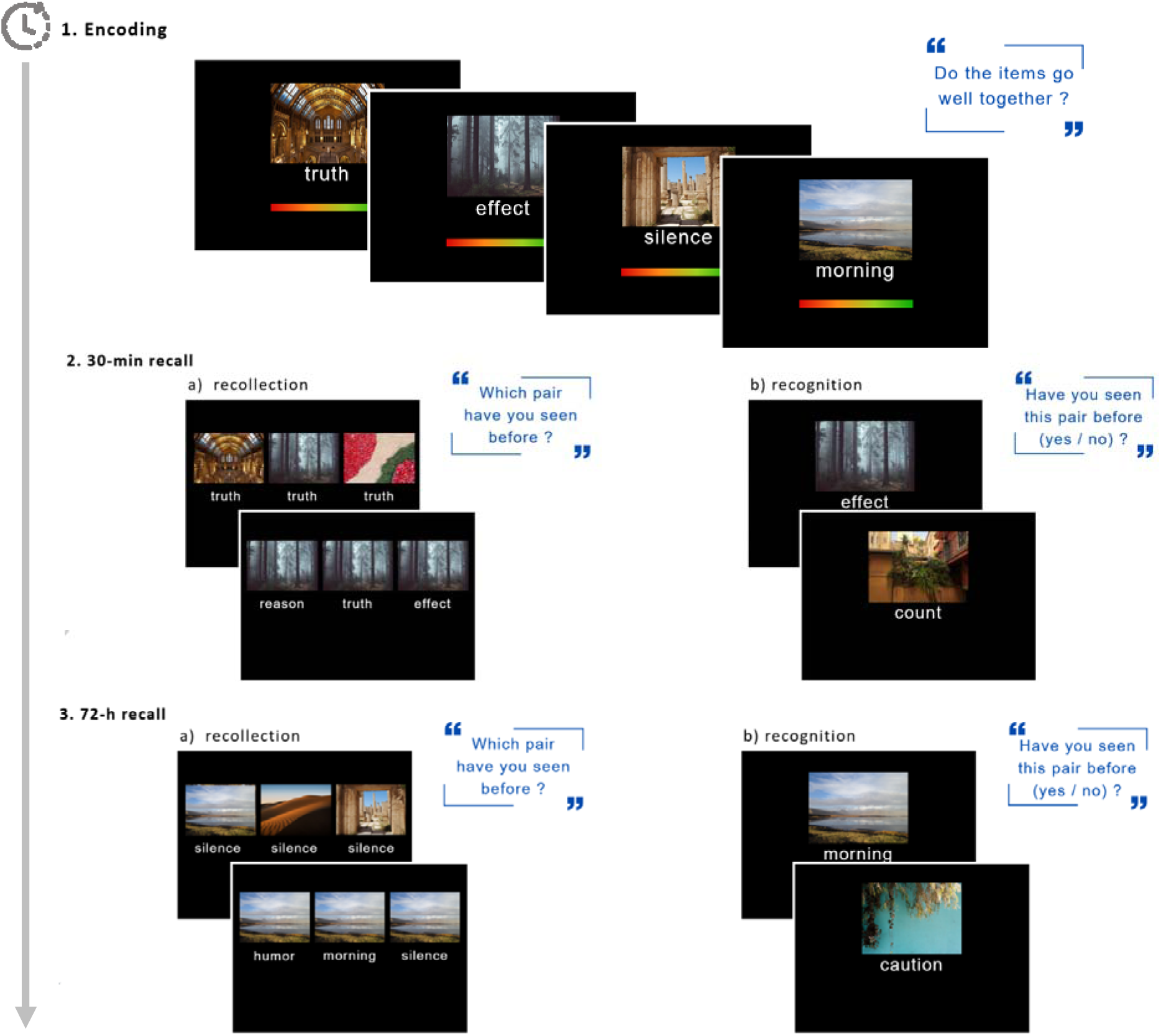
Illustration of the experimental associative memory task. **1 -** During the encoding, word-scene pairs were presented one at a time to participants who were asked to indicate how much the two items composing the pair were linked based on a subjective judgement. **2 -** 30 minutes after encoding, a first recall was proposed using half of the encoded pairs. This recall was performed by means of *a)* a recollection task during which the participants had to indicate which word had been presented with which scene during encoding or vice versa, followed by *b)* a recognition task during which they had to indicate whether or not the pairs presented sequentially had already been encountered during encoding. **3 -** 72 hours after the first session, a last recall using the remaining half of the encoded pairs was proposed according to the same procedure as the 30-minute recall.

#### 4.2 Stimuli

All stimuli were carefully selected according to the following parameters: no emotional salience, homogeneous word length and lexical frequency, balanced type of scenes’ environment (e.g., urban, rural, coastal, etc.). Beyond following one of the criteria proposed by Elliott and colleagues^16^ for measuring ALF, the use of both verbal and non-verbal stimuli was motivated by the need to employ multimodal material. It also aligned with the modality-based approach that traditionally guides the interpretation of cognitive profiles in PWE. We therefore used abstract words that were not conducive to visual imagery and photos of complex and rich visual scenes that could not be described in a univocal manner. The 56 to-be-remembered pairs presented during the encoding were generated from a fixed set of stimuli (56 abstract words, 56 scenes) according to 4 pseudo-randomised sets of pairs. Within each group (PWE vs. HC), participants were allocated throughout the inclusions to one of the 4 shuffles, i.e., 4 possible sets of pairs. By this mean, each participant was exposed to the same words and scenes, but not to the same pairing or presentation sequence. The methodology was identical for creating all the distractor pairs later used during the recall sessions.

#### 4.3 Experimental procedure

##### 4.3.1 Encoding

During the encoding session, the 56 pairs were successively presented on a computer screen. Participants were instructed in advance that they would later be asked to remember these pairs. However, they were informed that memorising all of them intentionally was considered unfeasible given the large number of stimuli. Each pair was displayed for a maximum exposure time of 5000ms.

At each trial, the participants were asked to provide a judgement about the congruence between the two stimuli. Using a methodology adapted from Borders and colleagues^33^, participants were thus instructed to rate, for each pair, the strength of the connection they personally made between the word and the scene on a 0 *(not at all congruent)* to 3 *(very much congruent)* coloured scale (Figure 1). This congruence judgement was introduced to promote the formation of an association between the two stimuli, thereby integrating a subjective perspective, as no correct or incorrect response was expected.

##### 4.3.2 Recollection

Memory performance was assessed 30min and 72h after the encoding session. At each time point, retrieval was first carried out using a recollection test designed to trigger the reconstruction of the original pairs, thereby engaging hippocampal-dependent associative processes and addressing the primary objective of this study. To this end, one stimulus presented during encoding (a word in 50% of trials and a scene in the remaining 50%) appeared on the screen for 500ms and was then associated for a further 4500ms together with 3 other stimuli: one target and two distractors. Participants were asked to select the pair presented during encoding, that is to indicate which word had been paired with which scene, or vice versa. The two distractors were categorised as (i) “familiar but not paired”, meaning that it had been encountered previously but in a different pair (i.e., Old/Old), or (ii) “hybrid”, meaning that it had never been encountered before (i.e., Old/New). Thus, the correct answer could not be based solely on familiarity since both the target and one of the distractors were presented at encoding.

Accordingly, we collected for the recollection task at both delays (i) the number of correct answers (CAs) out of 28 items, (ii) the number of false alarms (FA), (iii) the nature of the FA depending on the two types of distractors (i.e., familiar but not paired [FnP] or hybrid [H]), (iv) the number of non-responses (i.e., missing or delayed responses).

##### 4.3.3 Recognition

In line with recommendations for measuring ALF^16^, a recognition test was also performed. At both 30-min and 72-h delays, following the recollection test, a total of 56 pairs (28 targets, 28 distractors) were successively presented to the participants at a maximum rate of 4 seconds per pair. The instruction was to indicate whether the presented pair was one of the pairs seen during encoding. The 28 distractors were classified into 3 categories: (i) “familiar but not paired” (n=7) where both the scene and the word were seen but not paired; (ii) “hybrid” (n=14) where only the word (n=7) or the scene (n=7) was encountered at encoding but was here presented with a new stimulus; (iii) “new” (n=7) where both the scene and the word were new.

Accordingly, we collected for the recognition task both at 30min and 72h (i) the number of CAs out of 56 items, combining here the number of hits and the number of correct rejections, (ii) the number of FA, (iii) the nature of the FA depending on the three types of distractors (i.e., FnP, H or new [N]), (iv) the number of misses, (v) the number of non-responses (i.e., missing or delayed responses).

### 5. Outcome measures

As indicated above, the present analysis focuses on recollection-related outcomes, whereas data collected during recognition will be presented in a dedicated analysis. Accordingly, the primary endpoint of the present study was the recollection-based memory loss 72 hours after encoding in PWE compared to HC, defined as the evolution of CAs between 30-min and 72-h recollection.

Secondary endpoints related to recollection included comparisons between PWE and HC regarding (i) the qualitative distribution of errors (number and type of FA during recollection) at 30min and 72h; (ii) the relationship between the recollection performance at 30min and 72h and the nature of the congruence judgement made during encoding (proportion of FA relating to pairs that were judged *not at all* or not *very* congruent at encoding). Additional secondary endpoints specific to PWE explored the relationship between memory loss and clinical variables including (i) lobar localisation of the EZ (temporal vs. extra-temporal); (ii) duration of the disease; (iii) hippocampal sclerosis; (iv) seizure frequency; (iv) number of ASM.

### 6. Data analysis

#### 6.1 Sample size

A total of 80 participants (40 PWE and 40 HC) provided sufficient power to detect a memory loss at 72h twice as large as at 30min in PWE, assuming 10% memory loss in HC. Power was estimated via simulation using a Bernoulli distribution at the individual level (28 items; p = 0.8 at 30min for both groups, 0.7 at 72h for HC, and 0.6 at 72h for PWE) and a Wald test with a two-sided α = 0.05 on the group coefficient in a linear regression model of memory loss (number of CAs at 30min – number of CAs at 72h), adjusted for age, sex, and years of education. Covariate distributions were based on demographic data from PWE managed at the adult Epilepsy Department of Hospices Civils de Lyon between 2017 and 2019 who met the study eligibility criteria. The resulting power was 95% over 1000 simulations.

#### 6.2 Statistics

All analyses were performed using R, version 4.4.0.

Mean value of congruence judgement at encoding (considering that a HD value is equal to 1.5a value of 1.5 when no congruence decision could be made) was compared between PWE and HC using a test of Student.

Recollection data were analysed using a binary criterion for each trial: correct vs. incorrect answers, with non-responses and FA combined into the latter category. To confirm that participants performed above chance at each time point (30min and 72h), a logistic mixed-effects model was adjusted for each delay, including a random intercept for participants. At each time point, the model intercept was compared with −0.69, corresponding to the expected value under chance-level performance. To confirm that participants performed above chance at each time point (30min and 72h), a logistic mixed-effects model was adjusted for each delay, including a random intercept for participants. At each time point, the model intercept was compared with −0.69, corresponding to the expected value under chance-level performance

The main model was a logistic mixed-effects model including a linear effect of age at inclusion (centred on 38 years old), years of education (centred on 15 years), allocated shuffle (4 categories), group (PWE vs HC), time (30min vs 72h), as well as a group x time interaction and a random subject effect on the intercept. The relationship between the congruence judgement at encoding and the recollection performance at both delays was examined by adding congruence as a predictor, with a value of 1.5 assigned where no congruence decision could be made by the participant during encoding. Different effects of congruence according to time and group were tested by adding successively two (congruence x time and congruence x group) and three-way (congruence x time x group) interactions to the model. Among these models, the final one was chosen considering the significance of Wald’s test of each coefficient. The Type I error rate was set at 5% for Wald tests performed on model coefficients, except for tests performed on interaction coefficients, for which it was set at 10%.

For the analyses of epilepsy-related variables within the patient group, the main model was a logistic mixed-effects model including, as previously described, a linear effect of age at inclusion (centred on 38 years old), years of education (centred on 15 years), allocated shuffle (4 categories) and time (30min vs 72h). Localisation of epilepsy, number of ASM and seizure frequency were examined by adding the corresponding fixed effect and its interaction with time to the main model. Odds-ratio (OR) of the different variables are presented with their 95% confidence interval (CI).

## RESULTS

### 1. Population

A total of 41 PWE and 42 HC were recruited. One PWE withdrew consent before completing the cognitive task and two HC were excluded due to significant depressive and anxiety symptoms identified during their clinical evaluation. Accordingly, 40 PWE and 40 HC were included in the final analyses.

#### 1.1. Demographic and clinical characteristics

PWE and HC did not differ in age, sex ratio or years of education (Table 1). Thirty-five PWE (88%) and 34 HC (85%) were right-handed, 4 (10%) in each group were left-handed, 1 PWE and 2 HC were ambidextrous. According to the inclusion criteria, all PWE demonstrated normal results on a comprehensive neuropsychological assessment. However, 35 (87.5%) reported subjective memory complaints consistent with ALF.

**Table 1.**
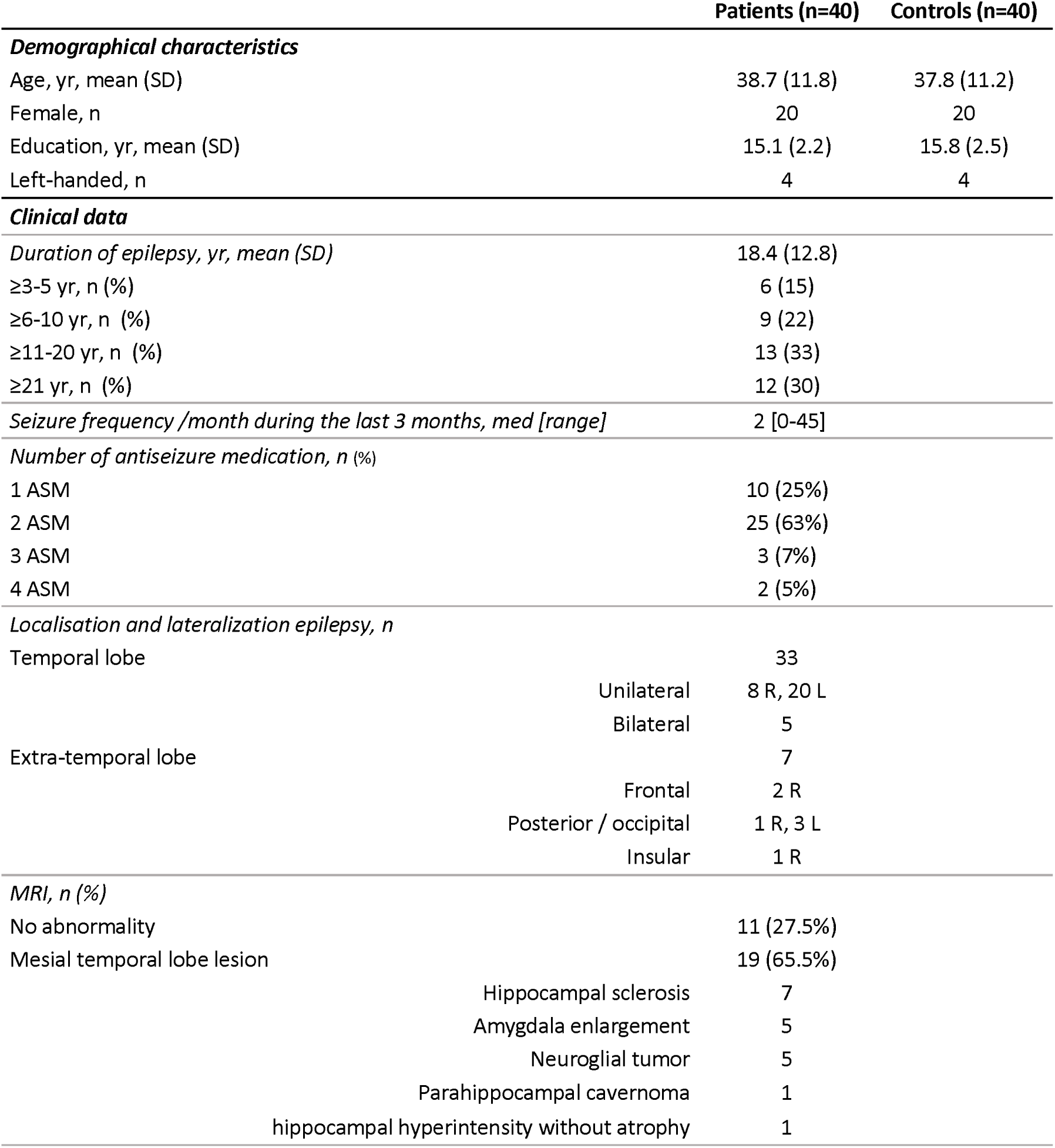
Participants’ demographic and clinical characteristics.

#### 1.2. Epilepsy-related features

The mean ± SD duration of epilepsy was 18.4 ± 12.8 years [range 3-53]. The median seizure frequency within the three months prior to inclusion was 2 seizures per month [range 0-45]. Only 3 patients reported focal to bilateral tonic-clonic seizures.

Overall, 33 PWE suffered from TLE and seven from extra-temporal lobe epilepsy. Epilepsy was lateralised to the right side in 12 (30%) patients, including eight with TLE, and to the left side in 23 (55%), including 20 with TLE. The five remaining patients (15%) suffered from bilateral TLE. Brain MRI was normal in 11 patients (27.5%), including eight (24.2%) with TLE, but revealed a structural abnormality in 29 patients. Specifically, 19 patients had a mesial temporal lobe lesion.

### 2. Cognitive and behavioural data

#### 2.1. Testing conditions

All PWE and HC completed the full experimental procedure including encoding, 30-min and 72-hour recollection and recognition tests. None of the participants reported adverse events and/or any unexpected life event during the 72-hour period of the study. No patient demonstrated seizure during the task or between the encoding phase and the 30-min recall. Eight patients reported at least one focal seizure in the interval between the 30-min and the 72-hour recall, but none used rescue therapy with benzodiazepines. No focal to bilateral tonic-clonic seizure was reported.

The distribution of the participants across the 4 shuffles was the following: 23 (28.7%) in shuffle 1 (11 HC and 12 PWE) and 19 (23.8%) in each of the shuffles 2, 3 and 4 (10 HC and 9 PWE in shuffles 2 and 3, 9 HC and 10 PWE in shuffle 4).

#### 2.2. Encoding

As shown in Figure 2, the congruence judgments were similar in HC and PWE. Mean value of congruence was equal to 1.21 for PWE and to 1.27 for HC (*p*=0.5).

**Figure 2.**
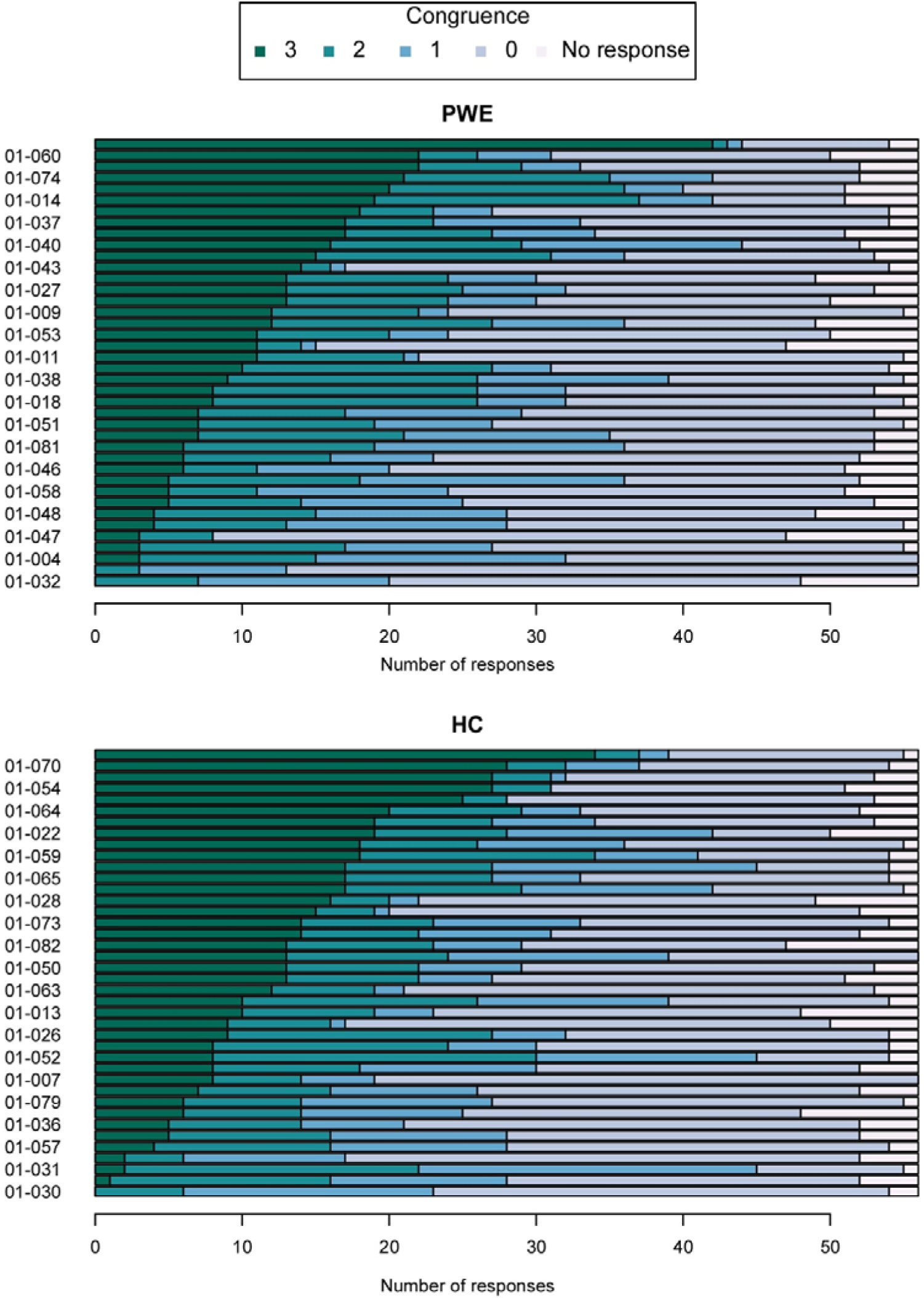
Distribution of congruence judgements at encoding. Each row represents an individual participant (PWE, top panel and HC bottom panel), and each coloured segment corresponds to the number of congruence judgement provided during encoding (0 = very weak congruence; 3 = very strong congruence). White segments indicate trials that result in non-responses.

#### 2.3. Recollection task

Analyses showed that all participants performed above chance on the recollection task at both 30min and 72h as each intercept was statistically different from −0.69, intercept estimation [95%CI] being respectively 1.23 [1.03;1.43] and −0.18 [-0.29;-0.07].

##### 2.3.1. Recollection score and memory loss

PWE demonstrated lower raw scores than HC at both delays. At 30min, the mean number ± SD of CAs was 19.08 ± 4.65 in PWE and 22.85 ± 3.24 in HC); at 72h, 11.60 ± 3.33 and 13.95 ± 3.19 respectively.

The logistic mixed-effects model revealed a significant memory decline 72h after encoding (Figure 3), with a lower recollection score at 72h than at 30min in both groups (PWE: OR [95%CI]: 0.31 [0.26;0.38]; HC: OR [95%CI]: 0.21 [0.17;0.26]). The memory loss between the two delays was greater in HC than in PWE. However, at 30-min recollection, PWE were less likely to perform correctly than HC (OR [95%CI]: 0.49 [0.38;0.63]) and, despite a smaller memory loss between 30min and 72h, PWE continued to perform significantly worse than HC at 72h (OR [95%CI]: 0.73 [0.58;0.91]).

**Figure 3:**
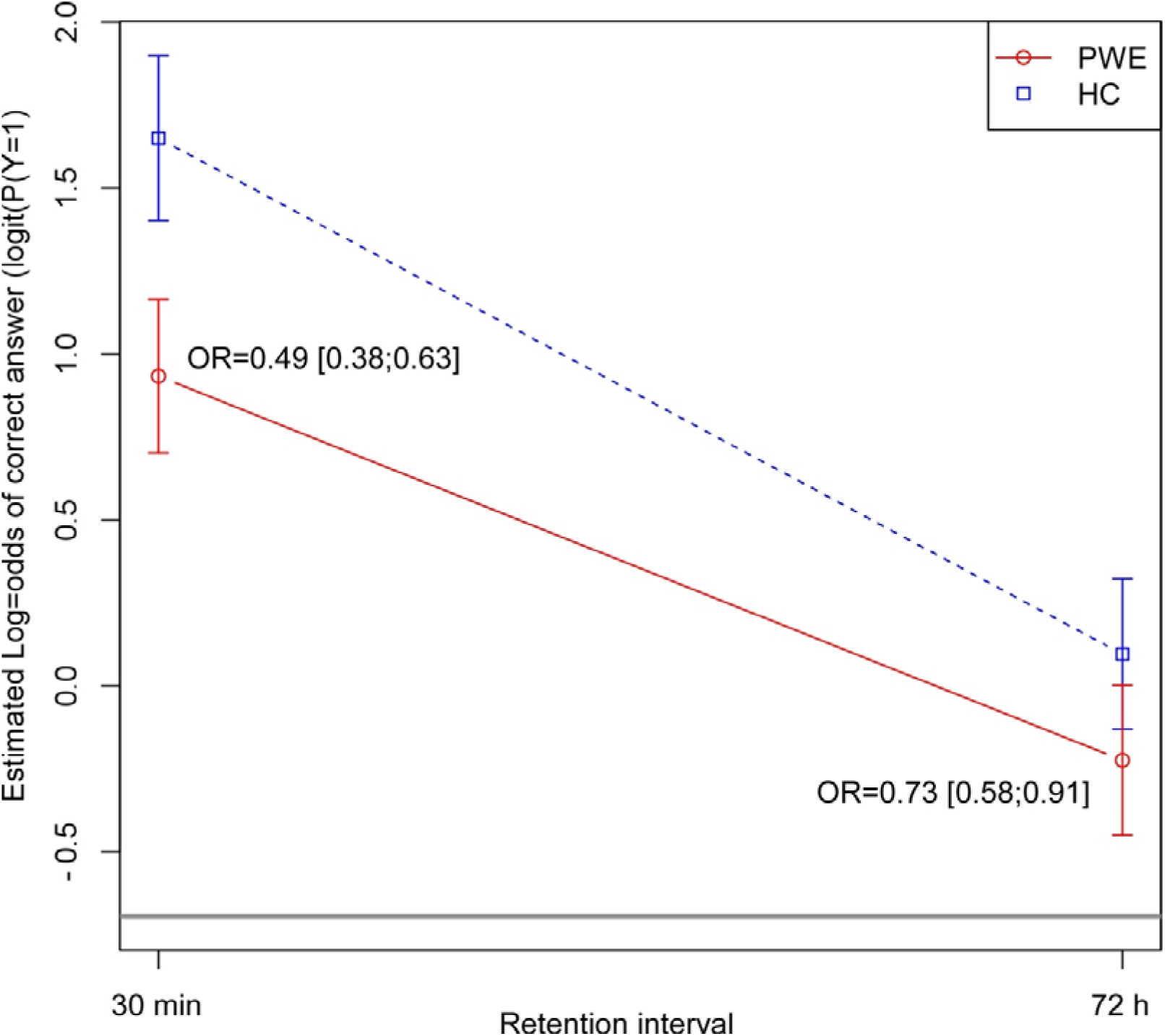
Temporal dynamics of recollection performance in patients with epilepsy (PWE) and healthy controls (HC) Estimated log-odds of a correct answer (logit(P(Y=1)) derived from the logistic mixed-effects model are shown for PWE (red solid line) and HC (blue dashed line) at 30min and 72h after encoding. Error bars represent 95% confidence intervals. Odds ratios (OR) for between-group comparisons are indicated at each time point. The horizontal grey line at the bottom of the figure indicates the chance-level threshold.

Concerning the effects of the others variables included in the logistic mixed-effects model, memory decline increased with age (OR [95%CI]: 0.98 [0.97;0.99]) but was not associated with years of education (OR [95%CI]: 1.04 [0.99;1.09]).

##### 2.3.2. Type of errors

We observed differences in the types of errors across groups and delays (Figure 4). The number of non-responses was higher in PWE (3.20 ± 2.64) than in HC (1.40 ± 1.15) at 30min, this difference was smaller at 72h (3.03 ± 2.13 for PWE; 2.08 ± 2.34 for HC). At 30min, the number of H-related FA was lowest in HC (0.65 ± 0.95) and lower than in PWE (1.38 ± 1.31). At 72h, this number increased in both groups, reaching 1.48 ± 1.24 in HC and 2.15 ± 1.64 in PWE.

**Figure 4.**
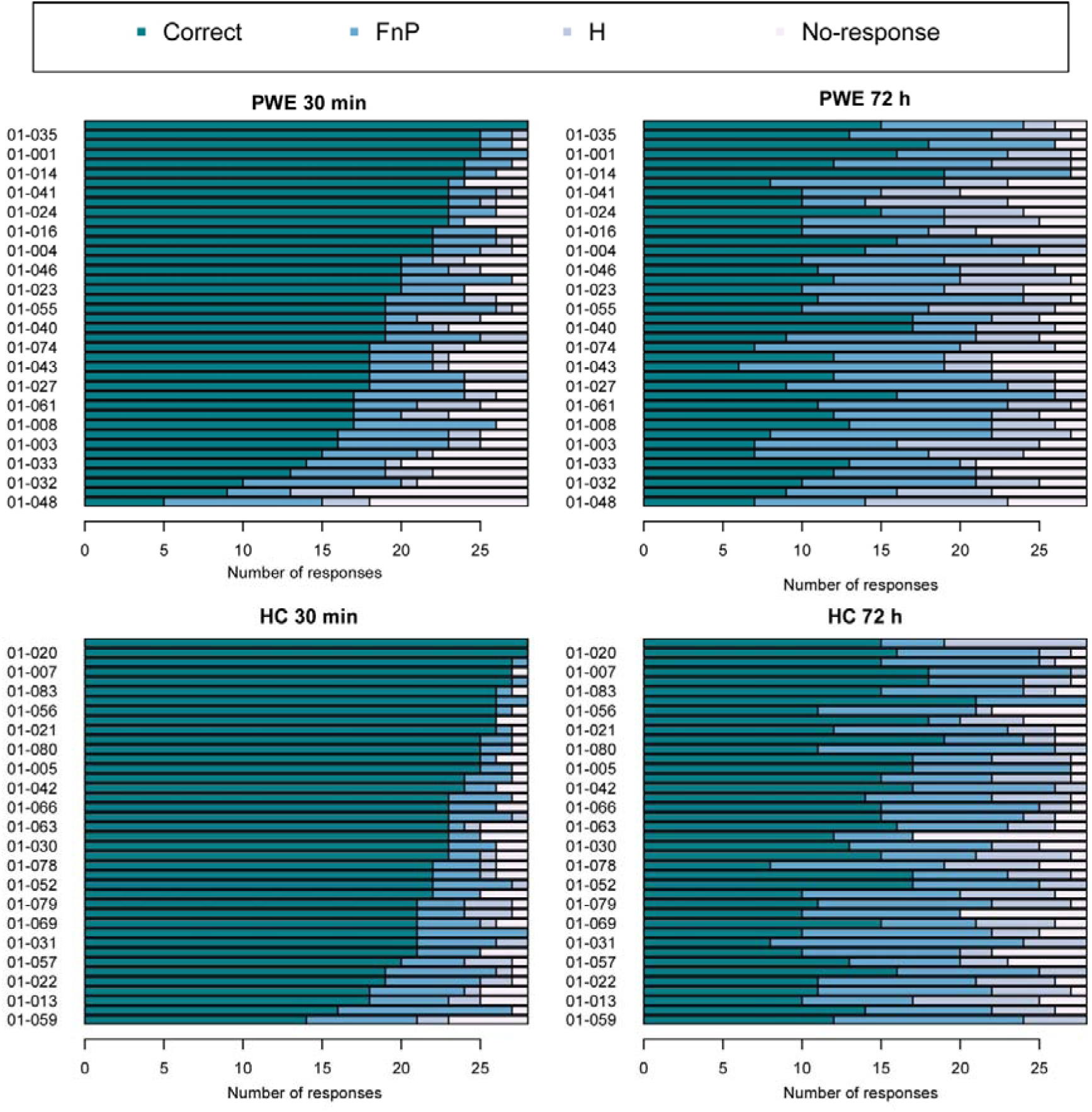
Distribution of type of responses in recollection at 30min (left panel) and 72h (right panel), in patients (PWE, top panel) and healthy controls (HC, bottom panel). Each row represents an individual participant, and each coloured segment corresponds to the number of responses per category (Correct answers; FnP-related false alarms, H-related false alarms). White segments indicate trials that resulted in non-responses.

##### 2.3.3. Effect of congruence judgement at encoding on recollection performance

Irrespective of group, congruence level at encoding (i.e., the strength of the association between the word and the scene) significantly affected recollection performance at 30min and 72h (p=0.03), indicating that congruence judgements at encoding influenced the subsequent memory performance. Specifically, the greater the level of congruence at encoding, the higher the subsequent recollection score. Moreover, irrespective of congruence level, time affected memory performance in both groups, with higher recollection scores at 30min than at 72h. At both delays, HC significantly outperformed PWE. However, as shown in Figure 5, the pattern of this effect differed between PWE and HC. A significant three-way interaction between congruence, time and group was thus observed (p=0.03). While the positive effect of congruence on recollection score was present both at 30min and 72h in PWE, it was only observed at 72h in HC.

**Figure 5.**
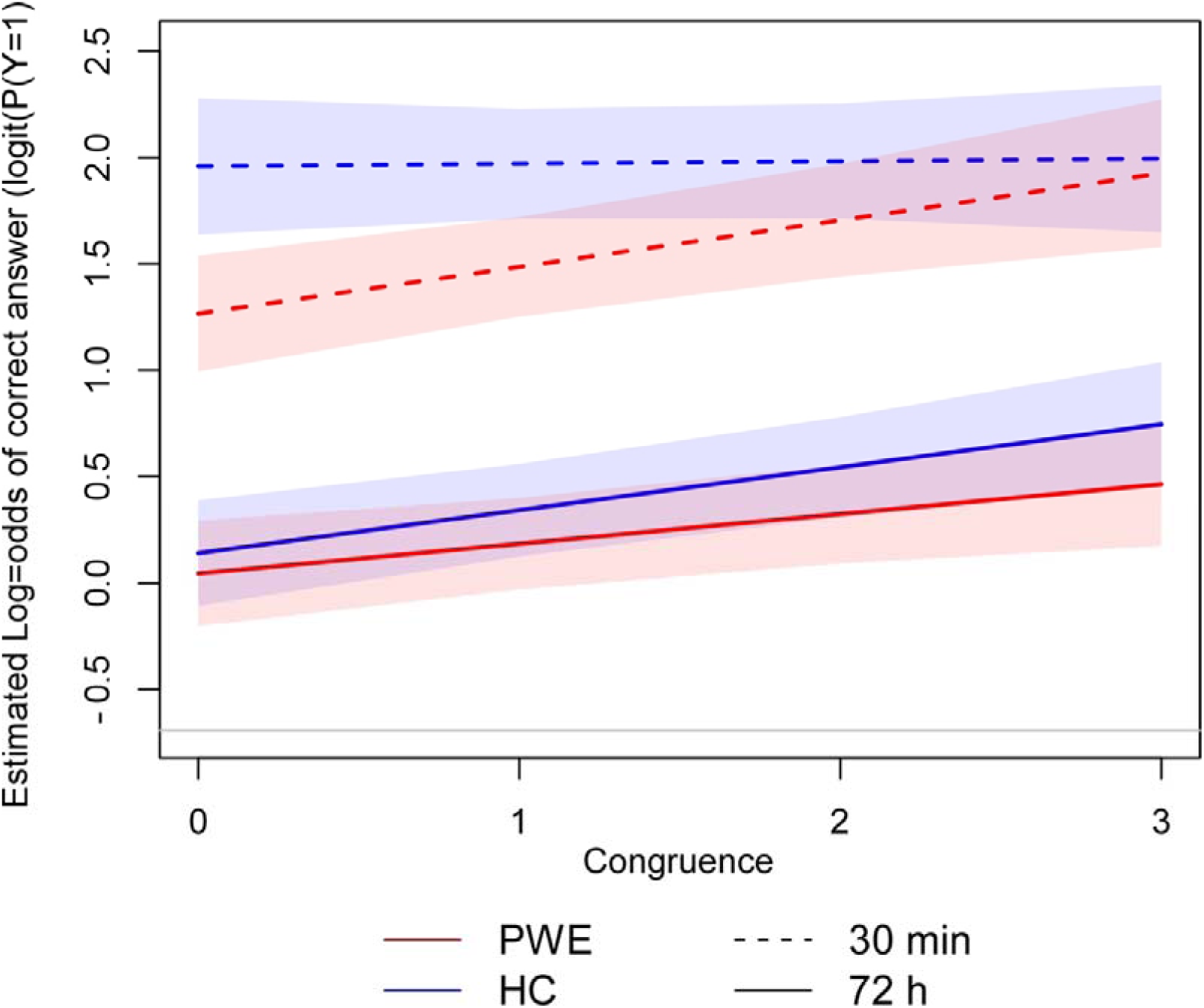
Differential effect of congruence judgement at encoding on recollection performance over time in patients with epilepsy (PWE) and healthy controls (HC)

##### 2.3.4. Relation between epilepsy features and memory decline in PWE

Memory loss at 72h did not significantly differ depending on the lobar localisation of epilepsy, i.e., TLE vs extra-TLE (at 72h OR [95%CI]: 0.68 [0.42;1.10]). It was not influenced by the number of ASM (OR [95%CI]: 0.95 [0.75;1.22]), the duration of epilepsy (OR [95%CI]: 1.00 [0.99;1.02]), nor the presence of a hippocampal sclerosis on MRI (OR [95%CI]: 1.02 [0.54;1.97]. No association was observed between memory loss at 72h and seizure frequency over the three months prior to inclusion (OR [95%CI]: 1.00 [0.99;1.02]).

## DISCUSSION

Both PWE and HC performed above chance at 30-min and 72-h recollection tasks, showing the potential methodological relevance of this associative memory paradigm in clinical context. Despite normal performance on a comprehensive neuropsychological assessment, a group difference emerged as early as 30min in our task, with significantly lower recollection scores in PWE than in HC. At this earlier delay, recollection in PWE appeared selectively supported by strong word-scene associations formed at encoding, whereas HC recalled congruent and non-congruent pairs equally well, suggesting efficient recollection regardless of associative strength. Memory loss over time appeared less pronounced than expected in PWE, despite persistently poorer performance at 72h. At this later delay, stronger word-scene associations enhanced recollection in both groups, indicating significant congruence effect.

While ALF is commonly interpreted as a disruption of active consolidation, it remains debated whether it results from a late disturbance of this process or a subtle deficit arising at the earliest stages of memory formation^19^. The group difference observed at 30-min recollection, more closely aligned with the latter hypothesis, shows that alterations in recollection can be detected in patients who otherwise perform normally on standardised memory tests, despite 87.5% reporting impeding complaints. Several studies using refined cognitive paradigms that target hippocampal-dependent mechanisms, such as novelty preference^34^, pattern separation^35^, or face-name associations^36^, have reported early memory impairments in PWE or neurodegenerative diseases, sometimes with no abnormal forgetting at longer delays^15,19,37–39^. Although assessment tools incorporating longer retention intervals or ecological approaches are valuable for assessing episodic memory^3^, tasks tapping into its associative dimension seem particularly complementary and informative for clinical purpose. This is especially relevant in TLE presurgical evaluation, where hippocampal function constitutes a critical concern. Moreover, such sensitive paradigms are easy to implement in clinical practice, as they do not require additional visits and are generally perceived as engaging by patients.

Beyond providing strong arguments regarding the added value of associative material for detecting episodic memory impairment in PWE, the significant group effect found at 30-min recollection raises questions about the origins of this apparent early vulnerability in memory formation among patients. Several non-mutually exclusive hypotheses might be to consider, including increased susceptibility to interference during encoding or early consolidation^40^, aberrant prediction errors during encoding^41^, disrupted hippocampally driven viewing behaviours^42^, or altered hippocampal-neocortical communication^43^. In all cases, the diversity of neurological contexts in which ALF has been reported further encourages the search for shared mechanistic explanations for possible impaired memory construction. In this regard, a recent review by Arulchelvan and Vanneste^44^ has proposed to conceptualise ALF not merely as a consolidation deficit, but as a maladaptative overactivation of forgetting mechanisms, potentially related to alterations in synaptic regulation and remodelling. Together with existing electrophysiological evidence highlighting the crucial role of cross-frequency coupling in memory consolidation^45^, this perspective opens new avenues for understanding the mechanisms underlying ALF, including beyond epilepsy.

The three-way congruence x time x group interaction further clarifies the between-group difference observed at 30-min recollection. Thus, PWE appeared able to memorise intermodal information as long as they established meaningful associations at encoding, probably engaging richer memory representations. Conversely, information that failed to elicit such associative processing appeared more vulnerable to forgetting, presumably due to shallower encoding. Consistent with this interpretation, one of the few studies that have used associative material to investigate ALF in patients with TLE found no evidence of ALF for encoded associations, indicating preserved recall of successfully formed associations in both patients and controls^46^. More broadly, multimodal encoding contexts have long been shown to facilitate memory formation and retention^47^. Future studies combining associative memory paradigms with direct neurophysiological measures will be critical to further elucidate the mechanisms underlying encoding and early consolidation in PWE, which appear central in our task.

On a behavioural level, the modulatory role of congruence on subsequent recall suggests that PWE could benefit from strategies promoting elaborative encoding, which is thought to be more effective than simple rehearsal^48^ but also transferable to everyday life. Training PWE to generate stronger, meaningful associations during encoding could therefore be incorporated into therapeutic education programs to foster compensatory strategies enhancing episodic memory, thereby alleviating the negative impact of memory impairment on patients’ quality of life.

Some limitations should be acknowledged, including the fact that the paradigm included a demanding 72-h delay without a short-delay baseline, which may restrict its applicability. Moreover, although no association was found between epilepsy-related features and memory loss over time, the clinical heterogeneity of the cohort in terms of epilepsy duration, seizure frequency and MRI lesions should be considered when interpreting the generalisability of our findings. Taken together, and within these constraints, this study nevertheless supports that ALF may, in some cases, reflect early consolidation abnormalities, consistent with a dynamic conception of memory consolidation. Accordingly, it encourages to prioritise associative materials when assessing episodic memory, but also to systematically consider the depth and quality of encoding. Continued efforts to better characterise memory complaints in PWE, particularly when suggestive of ALF, are essential to improve clinical care, reduce stigma and provide clinicians with appropriate tools to communicate effectively with patients and their relatives.

## Data Availability

All data produced in the present study are available upon reasonable request to the authors

## ACKNOWLEDGEMENT

We are grateful to Dr S. Hennion, Dr G. Plancher and Dr O. Bertrand for their suggestions and constructive comments about the design of the memory task and the conduct of the study.

We also thank all the patients and control participants who took part in this research.

The EPIMNESIE study was funded by the French Ministry of Health (PHRC Interregional 2020).

## DISCLOSURE OF CONFLICTS OF INTEREST

No author has conflict of interest related to this study

The statistical analyses were conducted by Emmanuelle Dantony, Mikaïl Nourredine and Pr Pascal Roy, Department of Biostatistics.

## Trial registration

ClinicalTrials.gov NCT04924933

---

**Supplemental Table 1.** List of the tests used in the routine neuropsychological assessment. The tests are presented here by main cognitive domain. A minimum of four memory tests was administered to each patient (two in the verbal modality and two in the visuo-non-verbal modality). The shaded tests were not systematically administered.

| Domain | Test used |
| --- | --- |
| **Global reasoning** |  |
| *Wechsler Adult Intelligence Scale - Fourth edition (WAIS-IV)* |  |
| Verbal comprehension index | Similarities<br>Information<br>Vocabulary |
| Perceptual Organization | Block Design<br>Matrix reasoning and or Visual Puzzles |
| **Memory** |  |
| Verbal memory | 16-item Free and Cued Recall<br>Logical Memory (Wechsler Memory Scale - Fourth edition WMS-IV)<br>Verbal Paired Associates (WMS-IV) |
| Non-verbal memory | Rey-Osterrieth complex figure test (ROCF)<br>Faces recognition (WMS-III)<br>Brief Visuospatial Memory Test - Revised (BVM-T-R) |
| **Executive Functions and Attention** |  |
| Working and short-term memory | Digit span (WAIS-IV)<br>Paced Auditory Serial Addition Test (PASAT) |
| Processing speed | Coding (WAIS-IV)<br>Trail-Making Test A |
| Shifting | Trail-Making Test B |
| Inhibition | Stroop test |
| Planification | "Test des commissions" |
| **Perception** |  |
|  | Visual Object Spatial Perception (VOSP) |
| **Language** |  |
| Visual naming | Boston Naming Test (BNT) or DO80 |
| Phonological fluency | Letter P |
| Semantic fluency | Category Animal |
| **Social cognition** |  |
|  | Emotion recognition (mini-SEA) |
| **Quality of life** |  |
|  | Quality of Life Inventory in Epilepsy - 31 |
| **Depression and anxiety** |  |
|  | Beck Depression Inventory (BDI-II)<br>State-Trait Anxiety Inventory (STAI) |

## REFERENCES

1. Hoppe C, Elger CE, Helmstaedter C. Long-term memory impairment in patients with focal epilepsy: Long-Term Memory Impairment in Patients with Focal Epilepsy. Epilepsia. 2007;48:26–29. doi:10.1111/j.1528-1167.2007.01397.x

2. Fisher RS, Vickrey BG, Gibson P, et al. The impact of epilepsy from the patient’s perspective I. Descriptions and subjective perceptions. Epilepsy Res. 2000;41(1):39–51. doi:10.1016/S0920-1211(00)00126-1

3. Lemesle B, Barbeau EJ, Milongo Rigal E, et al. Hidden Objective Memory Deficits Behind Subjective Memory Complaints in Patients With Temporal Lobe Epilepsy. Neurology. 2022;98(8):e818–e828. doi:10.1212/WNL.0000000000013212

4. Hohmann L, Berger J, Kastell S, Holtkamp M. DomainDspecific relationships of subjective and objective cognition in epilepsy. Epilepsia. 2023;64(7):1887–1899. doi:10.1111/epi.17624

5. Fitzgerald Z. Accelerated long-term forgetting: A newly identified memory impairment in epilepsy. J Clin Neurosci. Published online 2013:6.

6. Mameniškienė R, Puteikis K, Jasionis A, Jatužis D. A Review of Accelerated Long-Term Forgetting in Epilepsy. Brain Sci. 2020;10(12):945. doi:10.3390/brainsci10120945

7. Geurts S, van der Werf SP, Kwa VIH, Kessels RPC. Accelerated long-term forgetting after TIA or minor stroke: A more sensitive measure for detecting subtle memory dysfunction? Cortex. 2019;110:150–156. doi:10.1016/j.cortex.2018.04.002

8. Studer M, Guggisberg AG, Gyger N, Gutbrod K, Henke K, Heinemann D. Accelerated long-term forgetting in patients with acquired brain injury. Brain Inj. 2024;38(5):377–389. doi:10.1080/02699052.2024.2311349

9. Lah S, Black C, Gascoigne MB, Gott C, Epps A, Parry L. Accelerated Long-Term Forgetting Is Not Epilepsy Specific: Evidence from Childhood Traumatic Brain Injury. J Neurotrauma. 2017;34(17):2536–2544. doi:10.1089/neu.2016.4872

10. Maeder J, Sandini C, Zöller D, et al. Long-term verbal memory deficit and associated hippocampal alterations in 22q11.2 deletion syndrome. Child Neuropsychol. 2020;26(3):289–311. doi:10.1080/09297049.2019.1657392

11. Weston PSJ, Nicholas JM, Henley SMD, et al. Accelerated long-term forgetting in presymptomatic autosomal dominant Alzheimer’s disease: a cross-sectional study. Lancet Neurol. 2018;17(2):123–132. doi:10.1016/S1474-4422(17)30434-9

12. Rodini M, De Simone MS, Caltagirone C, Carlesimo GA. Accelerated long-term forgetting in neurodegenerative disorders: A systematic review of the literature. Neurosci Biobehav Rev. 2022;141:104815. doi:10.1016/j.neubiorev.2022.104815

13. Miller LA, Mothakunnel A, Flanagan E, Nikpour A, Thayer Z. Accelerated Long Term Forgetting in patients with focal seizures: Incidence rate and contributing factors. Epilepsy Behav. 2017;72:108–113. doi:10.1016/j.yebeh.2017.04.039

14. Lambert I, TramoniDNegre E, Lagarde S, et al. Accelerated longDterm forgetting in focal epilepsy: Do interictal spikes during sleep matter? Epilepsia. 2021;62(3):563–569. doi:10.1111/epi.16823

15. Cassel A, Morris R, Koutroumanidis M, Kopelman M. Forgetting in temporal lobe epilepsy: When does it become accelerated? Cortex. 2016;78:70–84. doi:10.1016/j.cortex.2016.02.005

16. Elliott Gemma, Isaac CL, Muhlert N. Measuring forgetting: A critical review of accelerated long-term forgetting studies. Cortex. 2014;54:16–32. doi:10.1016/j.cortex.2014.02.001

17. Baddeley AD, Atkinson AL, Hitch GJ, Allen RJ. Detecting accelerated long-term forgetting: A problem and some solutions. Cortex. 2021;142:237–251. doi:10.1016/j.cortex.2021.03.038

18. Atherton KE, Filippini N, Zeman AZJ, Nobre AC, Butler CR. Encoding-related brain activity and accelerated forgetting in transient epileptic amnesia. Cortex. 2019;110:127–140. doi:10.1016/j.cortex.2018.04.015

19. Cassel A, Kopelman MD. Have we forgotten about forgetting? A critical review of ‘accelerated long-term forgetting’ in temporal lobe epilepsy. Cortex. 2019;110:141–149. doi:10.1016/j.cortex.2017.12.012

20. Brissart H, Planton M, Bilger M, et al. French neuropsychological procedure consensus in epilepsy surgery. Epilepsy Behav. 2019;100:106522. doi:10.1016/j.yebeh.2019.106522

21. Laguitton V, Boutin M, Brissart H, et al. Neuropsychological assessment in pediatric epilepsy surgery: A French procedure consensus. Rev Neurol (Paris). 2024;180(6):494–506. doi:10.1016/j.neurol.2023.08.019

22. Lech RK, Suchan B. The medial temporal lobe: Memory and beyond. Behav Brain Res. 2013;254:45–49. doi:10.1016/j.bbr.2013.06.009

23. Gilboa A. Remembering Our Past: Functional Neuroanatomy of Recollection of Recent and Very Remote Personal Events. Cereb Cortex. 2004;14(11):1214–1225. doi:10.1093/cercor/bhh082

24. Addis DR, Moscovitch M, Crawley AP, McAndrews MP. Recollective qualities modulate hippocampal activation during autobiographical memory retrieval. Hippocampus. 2004;14(6):752–762. doi:10.1002/hipo.10215

25. Sheldon S, Levine B. Same as it ever was: Vividness modulates the similarities and differences between the neural networks that support retrieving remote and recent autobiographical memories. NeuroImage. 2013;83:880–891. doi:10.1016/j.neuroimage.2013.06.082

26. Des Jarlais DC, Lyles C, Crepaz N, the TREND Group. Improving the Reporting Quality of Nonrandomized Evaluations of Behavioral and Public Health Interventions: The TREND Statement. Am J Public Health. 2004;94(3):361–366. doi:10.2105/AJPH.94.3.361

27. Kwan P, Arzimanoglou A, Berg AT, et al. Definition of drug resistant epilepsy: consensus proposal by the ad hoc Task Force of the ILAE Commission on Therapeutic Strategies. Epilepsia 2010;51:1069–77

28. Hermann B, Busch RM, Reyes A, et al. A user’s guide for the International Classification of Cognitive Disorders in Epilepsy. Epileptic Disord. 2024;26(5):567–580. doi:10.1002/epd2.20268

29. Micoulaud-Franchi JA, Barkate G, Trébuchon-Da Fonseca A, et al. One step closer to a global tool for rapid screening of major depression in epilepsy: Validation of the French NDDI-E. Epilepsy Behav. 2015;44:11–16. doi:10.1016/j.yebeh.2014.12.011

30. Nasreddine ZS, Phillips NA, Bédirian V, et al. The Montreal Cognitive Assessment, MoCA: A Brief Screening Tool For Mild Cognitive Impairment. J Am Geriatr Soc. 2005;53(4):695–699. doi:10.1111/j.1532-5415.2005.53221.x

31. Wechsler D. Wechsler Adult Intelligence Scale - Fourth Edition (WAIS-IV). Published online 2011.

32. Bocéréan C, Dupret E. A validation study of the Hospital Anxiety and Depression Scale (HADS) in a large sample of French employees. BMC Psychiatry. 2014;14(1):354. doi:10.1186/s12888-014-0354-0

33. Borders AA, Aly M, Parks CM, Yonelinas AP. The hippocampus is particularly important for building associations across stimulus domains. Neuropsychologia. 2017;99:335–342. doi:10.1016/j.neuropsychologia.2017.03.032

34. Leeman-Markowski BA, Martin SP, Hardstone R, Tam DM, Devinsky O, Meador KJ. Novelty preference assessed by eye tracking: A sensitive measure of impaired recognition memory in epilepsy. Epilepsy Behav. 2024;155:109749. doi:10.1016/j.yebeh.2024.109749

35. Madar AD, Pfammatter JA, Bordenave J, et al. Deficits in Behavioral and Neuronal Pattern Separation in Temporal Lobe Epilepsy. J Neurosci. 2021;41(46):9669–9686. doi:10.1523/JNEUROSCI.2439-20.2021

36. De Simone MS, Rodini M, De Tollis M, Fadda L, Caltagirone C, Carlesimo GA. The diagnostic usefulness of experimental memory tasks for detecting subjective cognitive decline: Preliminary results in an Italian sample. Neuropsychology. 2023:636–649.

37. Mayes AR, Hunkin NM, Isaac C, Muhlert N. Are there distinct forms of accelerated forgetting and, if so, why? Cortex. 2019;110:115–126. doi:10.1016/j.cortex.2018.04.005

38. Contador I, Sánchez A, Kopelman MD, González de la Aleja J, Ruisoto P. Accelerated forgetting in temporal lobe epilepsy: When does it occur? Cortex. 2021;141:190–200. doi:10.1016/j.cortex.2021.03.035

39. Contador I, Sánchez A, Kopelman MD, González de la Aleja J. Long-term forgetting in temporal lobe epilepsy: Is this phenomenon a norm? Epilepsy Behav. 2017;77:30–32. doi:10.1016/j.yebeh.2017.09.020

40. Yonelinas AP, Ranganath C, Ekstrom AD, Wiltgen BJ. A contextual binding theory of episodic memory: systems consolidation reconsidered. Nat Rev Neurosci. 2019;20(6):364–375. doi:10.1038/s41583-019-0150-4

41. Aitken F, Kok P. Hippocampal representations switch from errors to predictions during acquisition of predictive associations. Nat Commun. 2022;13(1):3294. doi:10.1038/s41467-022-31040-w

42. Voss JL, Bridge DJ, Cohen NJ, Walker JA. A Closer Look at the Hippocampus and Memory. Trends Cogn Sci. 2017;21(8):577–588. doi:10.1016/j.tics.2017.05.008

43. Staresina BP, Wimber M. A Neural Chronometry of Memory Recall. Trends Cogn Sci. 2019;23(12):1071–1085. doi:10.1016/j.tics.2019.09.011

44. Arulchelvan E. Accelerated long-term forgetting across neuropsychiatric disorders: A review of mechanisms and a neurobiological perspective of forgetting. Published online 2026.

45. Lega B, Burke J, Jacobs J, Kahana MJ. Slow-Theta-to-Gamma Phase–Amplitude Coupling in Human Hippocampus Supports the Formation of New Episodic Memories. Cereb Cortex. 2016;26(1):268–278. doi:10.1093/cercor/bhu232

46. Audrain S, McAndrews MP. Cognitive and functional correlates of accelerated long-term forgetting in temporal lobe epilepsy. Cortex. 2019;110:101–114. doi:10.1016/j.cortex.2018.03.022

47. Long NM, Kahana MJ. Successful memory formation is driven by contextual encoding in the core memory network. NeuroImage. 2015;119:332–337. doi:10.1016/j.neuroimage.2015.06.073

48. Jaeger A, Martins THPG, Rodrigues JPP, Muniz BFB, Da Silveira Fonseca ALS, De Oliveira Gonçalves A. The benefits of elaborative encoding over retrieval practice for associative learning. Mem Cognit. 2025;53(5):1592–1607. doi:10.3758/s13421-024-01671-z

